# Estimating population immunity without serological testing

**DOI:** 10.1101/2020.04.23.20076786

**Authors:** Andrew Lesniewski

## Abstract

We propose an approximate methodology for estimating the overall level of immunity against COVID-19 in a population that has been affected by the recent epidemic. The methodology relies on the currently available mortality data and utilizes the properties of the SIR model. We illustrate the application of the method by estimating the recent levels of immunity in 10 US states with highest case numbers of COVID-19.

---

The purpose of this brief note is to propose an approximate methodology for determining the level of immunity in a population affected by (an infectious disease such as) COVID-19. In the absence of reliable, large scale testing, paucity and spotty character of case data, our approach relies on the mortality data.

The proposed methodology is predicated on the following simplifying assumptions.

i. The deterministic SIR model [1] is an adequate mathematical specification of the COVID-19 epidemic. In particular, we assume that individuals who have recovered from an infection develop a long lasting immunity against reinfection.
ii. The basic reproduction ratio has the value given by

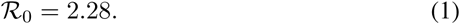 This value was found to be the midpoint of the range estimated by means of MLE in [8], see also [5].
iii. The infection rate parameter of the SIR model is given by

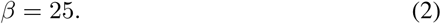 This choice is motivated by the fact that *τ* = 1*/β* is the characteristic time scale (in the units of a year) of the spread of the epidemic. Given that the length of the full cycle of COVID-19 appears to be about 6 months, we believe that any choice in the range of 20 to 30 would be appropriate. The choice above is probably conservative. The above values of *R*_0_ and *β* dictate the following value of the recovery rate parameter *γ* = *β/R*_0_ of the SIR model:

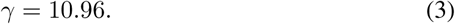
iv. The reliance on the mortality data introduces a lag of *l* days between the date when an individual contracts the disease and the date they die. For the estimates below we assume

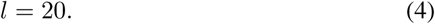 This choice is justified by a number of studies, see e.g. [2] and references therein: “The symptoms of COVID-19 infection appear after an incubation period of approximately 5.2 days. The period from the onset of COVID-19 symptoms to death ranged from 6 to 41 days with a median of 14 days”.
v. The infection fatality rate (IFR) is given by the value

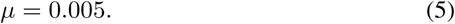

There is a wide spectrum of estimates of *µ* in the literature, see e.g. [3], [4], [6], based on complete tests conducted on small closed populations exposed to COVID-19, but there appears to be a consensus that the actual range should be between 0.1% and 1.0%. The difficulty lies of course in the fact that the IFR is based not only on the observed cases, but also on undiagnosed and asymptomatic cases. It also shows a strong dependence on other factors such as the patient’s age.

Notice that all parameter values selected above are population averages. Assuming distributions of values across risk factors such as incubation length, age group, geography, etc. could conceivably lead to more refined forecasts.

Our algorithm relies on the following properties of the SIR model. Let *S*(*t*), *I*(*t*), and *R*(*t*) denote the population fractions of susceptible, infected, and removed at time *t*, respectively. Note that, in this model, *R*(*t*) includes the individuals who have left the population through mortality at the rate *µ*. Then:

i. *S*(*t*), *I*(*t*), and *R*(*t*) add up to 1, for all *t*:

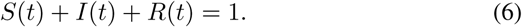
ii. The following conservation law holds for all *t*:

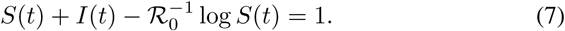
iii. The following relation holds:

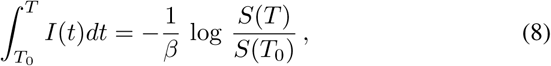

where *T*_0_ *< T*.

We now proceed as follows. We replace the continuous time variable *t* with daily samplings *t*_*n*_ = *nδ, n* = 0, 1, …, n, where *δ* = 1*/*365, and denote *R*_*n*_ *≡ R*(*t*_*n*_). Here, *t*_n_ = *T* corresponds to the latest observation, and *t*_n*−l*_ = *T*_*l*_ corresponds to the lagged date *T*_*l*_ = *T− l* days. We estimate *R*_n*−l*_, the removed fraction on the date n *− l*, as

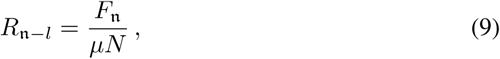

where *N* is the population size, and *F*_*n*_ is the total number of fatalities up to date *n*. Properties (i) and (ii) above imply that 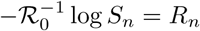, i.e.

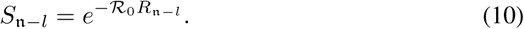

This implies that

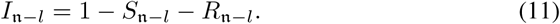

Now, from the third property we conclude that

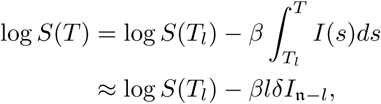

i.e.

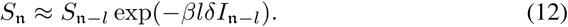

Similarly,

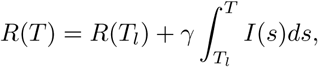

i.e.

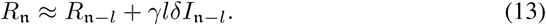

Also,

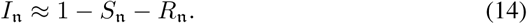

The approximations used in the calculation above are reasonable, but it is impossible to give an *a priori* estimate of the error made: they may equally well overshoot or undershoot the actual value of the integral 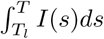. We should also point out that the algorithm is extremely easy to implement in computer code.

Assuming that all individuals who have recovered from the infection develop a long lasting immunity to the disease, we are thus led to the following estimator of the percentage of individuals with immunity:

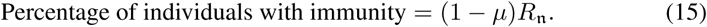

We have used the estimator (15) for the recent data of the 10 US states with the largest numbers on COVID-19 cases. The results, using the data taken from [7] as of April 20, April 21, and April 22 of 2020, are summarized in Table 1. Each of the columns contains the estimated percentages of immune individuals for the date indicated.

**Table 1:** Estimated percentages of populations immune against COVID-19

| State | % Imm 04-20 | % Imm 04-21 | % Imm 04-22 |
| --- | --- | --- | --- |
| NY | 29.16 | 30.15 | 31.00 |
| NJ | 15.95 | 17.20 | 18.22 |
| MA | 8.76 | 9.46 | 10.47 |
| PA | 3.64 | 4.34 | 4.60 |
| CA | 1.08 | 1.15 | 1.26 |
| MI | 8.33 | 9.08 | 9.44 |
| IL | 3.68 | 4.00 | 4.26 |
| FL | 1.34 | 1.41 | 1.51 |
| LA | 9.57 | 10.09 | 10.56 |
| TX | 0.61 | 0.64 | 0.67 |

It is possible that our estimates, due to the conservative assumptions made, might actually somewhat underestimate the actual levels of immunity. They do, however, show enough variability among the states that they might be of interest as rough estimates of the current levels of immunity against COVID-19.

## Data Availability

The data used is in public domain (Worldometer.com).

## Acknowledgement

I would like to thank Margaret MacNeil and Nicholas Lesniewski for discussions.

